# Predicting Donor Selection and Multi-Organ Transplantation within Organ Procurement Organizations Using Machine Learning

**DOI:** 10.1101/2024.02.03.24302297

**Authors:** Chelsea Tanchip, Mohammad Noaeen, Kamyar Kazari, Zahra Shakeri

## Abstract

Organ procurement organizations (OPOs) play a crucial role in the field of organ transplantation, serving as key intermediaries in the process of organ donation. However, despite their vital function, there exists a pressing issue of transparency within the organ allocation process. This opacity not only impedes the overall effectiveness of OPOs but also raises ethical and societal concerns regarding organ distribution. This study utilizes the recently published ORCHID dataset, containing 133,101 records of organ donor referrals, to understand organ procurement and donor selection strategies in OPOs using machine learning (ML). We developed seven ML classification models to predict donor selection and the likelihood of at least four organs being suitable for transplantation, in line with established definitions of multi-organ transplantation. The models demonstrated variable recall values for donor selection, ranging between 0.62 and 0.80, while achieving consistently high performance across other evaluation metrics, notably with AUC values exceeding 0.95. Particularly in the context of multi-organ transplant predictions, the models exhibited remarkable effectiveness, with recall values spanning from 0.88 to 0.98 and AUC metrics consistently above 0.97. Administrative milestones and particular organ transplants were identified as key determinants in the organ allocation process. This study’s findings suggest significant opportunities to improve organ allocation strategies by focusing on the optimization of administrative practices, highlighting their substantial impact on transplantation success rates.

## I. Introduction

Organ transplantation is a vital, life-saving procedure for individuals with end-stage diseases. Single-organ and multi-organ transplants (transplanting multiple organs from the same donor) have exponentially grown in demand over the past few years [1]. Yet, a significant disparity exists between the number of organ donors and those awaiting transplantation. In 2023, only 45% of 103,365 people on the organ transplant waiting list have actually received transplants in the USA [2]. This disparity is believed to stem from inequities related to social determinants of health (SDoH) and administrative factors, including transplant center characteristics [3, 4].

The organ procurement process typically occurs within acute-care hospitals, but the process has long been constrained by logistical and practical challenges. [5]. Organ procurement organizations (OPOs) were established to improve upon the challenges presented in hospitals, being dedicated solely to organ procurement. However, there is a lack of transparent data available from OPOs, which makes the decision-making process towards organ procurement and transplantation unclear.

Machine learning (ML) has become a widely adopted method for predicting organ allocation and donor-recipient suitability [6–8]. ML’s ability to detect new patterns from data lends itself well as a clinical decision-making aid. In this study, we aim to use ML primarily to understand and predict the clinical and administrative factors that influence donor selection and multi-organ transplantation in an OPO setting. This is the first study to analyze organ donor selection factors and multi-transplant outcomes in a multi-center OPO context. This study addresses the following research questions (RQs):

***RQ1:*** What factors influence the selection of a referred donor for organ transplantation?

***RQ2:*** What factors influence the event of multiple organs being transplanted from one donor after selection?

To address these RQs, we leverage the Organ Retrieval and Collection of Health Information for Donation (ORCHID) dataset to explore the factors influencing organ donor selection and the feasibility of multi-organ transplantation within OPOs, using advanced machine learning techniques. By identifying critical determinants in the donor selection process and evaluating their impact on multi-organ transplant outcomes, our research aims to contribute significantly to the optimization of organ allocation and transplantation practices. Through this analysis, we seek to address the pressing need for transparency and efficiency in the organ donation process, offering a data-driven approach to enhancing the effectiveness of OPO operations.

## II. Methods

### A. Data Collection and Preparation

#### Dataset

In July 2023, the Organ Retrieval and Collection of Health Information for Donation (ORCHID) was published [9]. The ORCHID dataset is the first multi-center dataset to be collected from deceased donor referrals across six different organ procurement organizations (OPOs) across the USA for six years, between January 1, 2015 and December 31, 2021. The ORCHID dataset contains 133,101 records of organ donor referrals and 34 variables related to clinical and administrative attributes associated with their referral. The administrative attributes include binary checkpoints for whether the donor passed different procedural milestones (e.g., referred to an OPO, approached) leading to selection for transplant. The dataset also contains columns for eight different organs (heart, liver, left and right kidney, left and right lung, pancreas, intestine). These columns’ values refer to the outcome of organ transplantation: whether that organ was ultimately transplanted, recovered for research, or recovered for transplant but ended up being used for research. The dataset was acquired from PhysioNet [10].

#### Data inclusion and exclusion

The full dataset was used for ***RQ1***. For ***RQ2***, in order to identify factors predictive of multi-organ transplantation, only donors who were selected for organ transplant (N = 8,972, 7% of dataset) were included for analysis.

#### Missing data

For variables missing at random (MAR), for example, *age, Cause_of_Death_OPO*), missing values were imputed using median for numerical variables and mode for categorical and ordinal variables. Variables missing not at random (MNAR), for instance, *timestamps* for administrative milestones were deliberately falsified and dropped. For the organ outcome variables, the data was considered MNAR as the outcome was none of the options (that specific organ was not recovered or transplanted, while at least one other organ was). The missing data for these specific variables is labeled as: *Not Recovered or Transplanted*.

#### Class imbalance

The full dataset is highly imbalanced, with 93% of potential donors not being selected for transplantation, while the remaining 7% were selected. SMOTE over-sampling [11] was applied to the minority class to account for the class imbalance in the training data. For the subset created in ***RQ2***, 5,503 (61%) individuals had less than four organs transplanted and 3,469 (39%) had at least four transplanted (out of eight organs total). Oversampling was not applied to this subset as the difference was not as stark as in ***RQ1*** and the subset was considerably smaller in size.

#### Feature engineering

For ***RQ1***, the specific organ outcomes were excluded since they are relevant to only ***RQ2***. The time-related variables were considered MNAR as they were derived from a specific event (e.g., *time_approached* is only filled out if the donor’s family was approached). In addition, the dataset creators randomly shifted all timestamps for privacy, rendering them inaccurate. For these reasons, all time-related variables were dropped for ***RQ1*** and ***RQ2***. However, new features were created based on the interval between these time variables (e.g., referral and approach), as this information was preserved by the dataset creators. The interval was grouped into three categories: whether less than 24 hours have passed between the variables, more than 24 hours, or the event never occurred. In each of the Cause of Death (OPO and UNOS), Mechanism of Death, and Circumstances of Death variable columns, values were grouped together based on the International Classification of Diseases 11th Revision (ICD-11) framework, the global standard for diagnostic health information. For ***RQ2***, the *transplanted* feature was dropped, since it is the inclusion criterion for membership in this subset (as everyone in this reduced set would already have transplanted as True). Building from the eight columns indicating transplantation outcomes for each organ type, a new binary feature was created: *multiple_transplants*, which denotes whether a) less than four organs were successfully transplanted, or b) at least four organs were successfully transplanted. The value of four or more organs transplanted signifying multi-organ transplantation has been used in previous studies [12].

### B. Model Development and Evaluation

Several models well-known for binary classification were developed: K-Nearest Neighbors (KNN), Logistic Regression (LR), Random Forest (RF), Support Vector Machine (SVM), Categorical Naï ve Bayes (NB), XGBoost, and Artificial Neural Network (ANN). The outcome variable was the binary *transplanted* variable in ***RQ1***, and *multiple transplants* in ***RQ2***. All other features that remained after pre-processing served as input variables. For each model, a nested stratified 5-fold cross-validation (CV) with grid search was implemented, which can find the optimal hyperparameters without producing an overly-optimistic CV score, as it involved conducting CV using the best hyperparameters determined through grid search. In each CV fold, the data was subject to imputation, standardization, and oversampling. Categorical variables were coded into dummy variables using one-hot encoding. Ordinal variables (e.g., day of the week referred) were encoded with label encoding to preserve their order. A held-out test set with 30% of the data was kept imbalanced (no oversampling applied) to maintain the real-world nature of the data. Missing data in the test set was imputed using the training set’s median of numerical variables and mode of categorical and ordinal variables. This method ensures that the test set remains an unbiased evaluation of the model’s real-world performance. The model was evaluated using the mean recall, precision, F1-score, and AUC score for each cross-validation fold (for the validation set) and the held-out test set (using the best hyperparameter combinations identified in CV). Classification results from the CV stage were compared between oversampled and the raw unbalanced data. Lastly, SHapley Additive exPlanations (SHAP) analysis [13] was implemented with the best-performing model per research question to quantify each feature’s impact on the model output. To ensure replicability and facilitate further research, the source code of all the presented machine learning models is available on GitHub^1^.

## III. Results and Discussion

### A. RQ1: What factors influence the selection of a referred donor for organ transplantation?

The input feature set was reduced to 16 features (8 clinical/demographic, 8 administrative) after pre-processing and feature engineering. Most notably, eye and tissue referral features were combined, while *brain_death, authorized, time_approached_to_authorized, procured*, and *time_authorized_to_procured* were dropped due to multi-collinearity with other features.

Table I shows cross-validation results and Table II shows the performance on the held-out test set, which revealed significantly different results for some models. Recall was significantly lower for LR, SVM, and NB (dropping from 0.81-0.99 in the CV stage to 0.58-0.80 in testing). In contrast, the precision values were moderately high in the CV stage (0.63-0.80) but improved considerably in testing (LR and SVM yielded 0.99). The AUC values in the testing set were similar to or even higher than the CV results for all models. Altogether, the disparity between some of the CV and held-out test results suggests overfitting in the LR, SVM, and NB models. Overfitting might have occurred because of the small amount of features after pre-processing, resulting in a simpler model. We attempted to avoid overfitting by removing multi-collinear features and applying regularization, which are valid techniques for models like LR and SVM. However, these techniques likely could not remedy the amount of information that might have been lost with feature reduction, given that the number of features in this dataset was already small to begin with. Nevertheless, XGBoost yielded the highest recall (0.80) and F1-score (0.83). The SHAP analysis of the XGBoost model revealed that the *approached* and *time_asystole_to_referral* (specifically if the referral milestone was not reached from after asystole) variables had the most impact.

**TABLE I:** Mean evaluation metrics from nested 5-fold cross-validation for raw unbalanced data and balanced data with SMOTE oversampling for donor selection prediction.

| Models | Recall |  | Precision |  | F1 |  | AUC |  |
| --- | --- | --- | --- | --- | --- | --- | --- | --- |
|  | Unbalanced | Balanced | Unbalanced | Balanced | Unbalanced | Balanced | Unbalanced | Balanced |
| KNN | 0.80 | **0.79** | 0.75 | **0.75** | 0.77 | **0.77** | 0.89 | **0.89** |
| LR | 0.81 | **0.99** | 0.78 | **0.59** | 0.79 | **0.74** | 0.90 | **0.97** |
| RF | 0.82 | **0.98** | 0.80 | **0.65** | 0.81 | **0.78** | 0.90 | **0.97** |
| SVM | 0.99 | **0.98** | 0.63 | **0.65** | 0.77 | **0.78** | 0.97 | **0.97** |
| NB | 0.94 | **0.97** | 0.60 | **0.56** | 0.74 | **0.71** | 0.95 | **0.96** |
| XGBoost | 0.85 | **0.88** | 0.80 | **0.78** | 0.82 | **0.83** | 0.92 | **0.93** |
| ANN | 0.83 | **0.95** | 0.79 | **0.68** | 0.81 | **0.79** | 0.91 | **0.96** |

**TABLE II:**
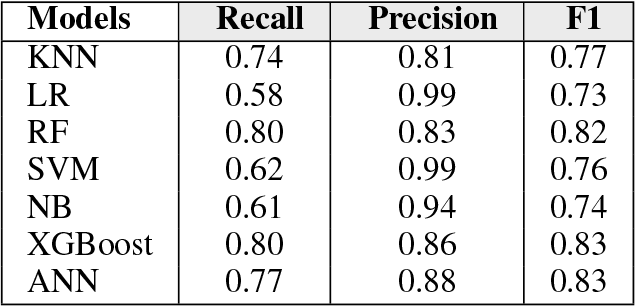
Comparison of performance metrics on the held-out test set for donor selection prediction.

Overall, all ML models were moderately effective in predicting donor selection within OPOs. Precision values were generally higher than recall, which was concerning since having many false negatives may hinder much-needed access to appropriate organ donors. The decrease in recall values between model development and testing suggested overfitting, which despite our efforts was unavoidable due to the limited size of the feature set. XGBoost showed consistently high results. The SHAP analysis (Fig. 1a) revealed that administrative milestones (*approached* (approaching the donor’s family) and *referred* (donor referral) not occurring after asystole are the most predictive of donor selection, which highlights the importance of administration and external communication in the organ procurement process. Obtaining family consent for organ donors is a known barrier to organ transplants [14], which in this study held more weight than the donor’s individual characteristics.

**Fig. 1:**
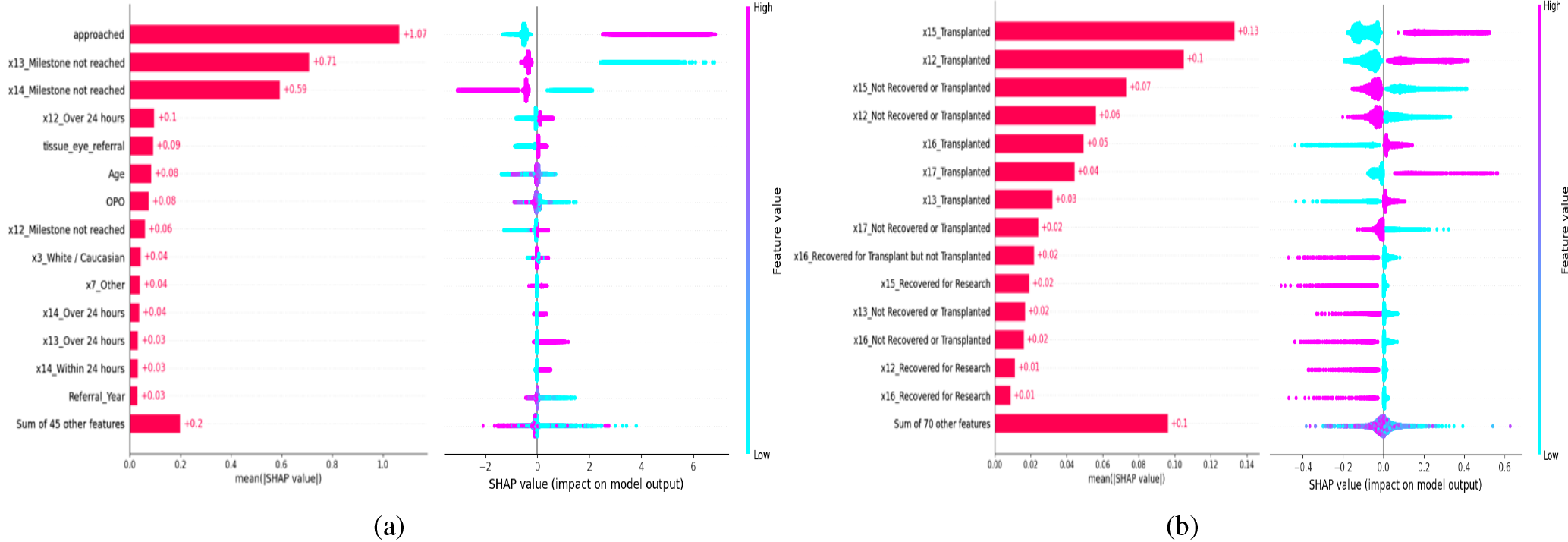
Top 15 SHAP feature importances for (a) XGBoost model predicting donor selection and (b) LR model for multi-organ transplantation. Feature labels in (a): x3 = *race*, x7 = *circumstances of death*, x12 = *time between asystole-referral*, x13 = *time between brain death-referral*, x14 = *time between referral-approach*. Feature labels in (b): x12 = *heart transplant outcome*, x13 = *liver transplant outcome*, x15 = *lung transplant outcome*, x16 = *kidney transplant outcome*, x17 = *pancreas transplant outcome*

### B. RQ2: What factors influence the event of multiple organs being transplanted from one donor after selection?

The input feature set included 25 features after preprocessing and feature engineering. Multi-collinearity was found between left and right kidneys, as well as left and right lungs, resulting in combining outcomes into *outcome_kidney* and *outcome_lung* respectively. Overall, classification results were excellent (Table III). LR, RF, XGBoost, and ANN in particular yielded consistently high (*≥* 0.97) results for recall, precision, and AUC.

**TABLE III:** Mean evaluation metrics from nested 5-fold cross-validation for multi-organ transplant prediction.

| Models | Recall | Precision | F1 | AUC |
| --- | --- | --- | --- | --- |
| KNN | 0.88 | 0.89 | 0.89 | 0.91 |
| LR | 0.98 | 0.98 | 0.98 | 0.99 |
| RF | 0.98 | 0.98 | 0.98 | 0.98 |
| SVM | 0.88 | 0.93 | 0.90 | 0.92 |
| NB | 0.91 | 0.86 | 0.88 | 0.91 |
| XGBoost | 0.97 | 0.98 | 0.98 | 0.97 |
| ANN | 0.98 | 0.98 | 0.98 | 0.98 |

Table IV shows that model performance on the held-out test set was similar to the results in the CV procedure, particularly LR, RF, XGBoost, and ANN, which produced metrics of at least 0.95. These models yielded consistently high precision and recall values in predicting multi-organ transplantation. LR, RF, ANN, and XGBoost yielded the highest AUC. SHAP was applied to the LR model as it yielded the highest metrics in all the categories (precision, recall, and AUC). The analysis (Fig. 1b) revealed that individual organ outcomes -those of particularly heart and lung transplants-were the most predictive of multi-organ transplantation.

**TABLE IV:** Comparison of performance metrics on the held-out test set for multi-organ transplant prediction.

| Models | Recall | Precision | F1 |
| --- | --- | --- | --- |
| KNN | 0.91 | 0.90 | 0.90 |
| LR | 0.98 | 0.99 | 0.98 |
| RF | 0.98 | 0.98 | 0.98 |
| SVM | 0.93 | 0.90 | 0.91 |
| NB | 0.88 | 0.91 | 0.90 |
| XGBoost | 0.98 | 0.95 | 0.97 |
| ANN | 0.97 | 0.99 | 0.97 |

Overall, the seven ML models were very effective in predicting multi-organ transplants from OPO-sourced donors. The models generally performed well and did not overfit. Successful transplantation of the heart and lungs was most likely to lead to more organs being transplanted from the same donor. These findings appear to support real-world organ allocation policies. For instance, the Organ Procurement and Transplantation Network (OPTN) policies state that when a candidate is eligible to receive a heart, lung, or liver, the second required organ would be allocated to the recipient from the same donor if the donor is located in the same local organ distribution unit where the recipient is registered [15]. The results of the present study may encourage the continued implementation of this policy in an effort to enhance organ allocation and equity.

## IV. Conclusion

There is a considerable lack of transparency in the decision-making process underlying donor transplant selection and organ outcomes within OPOs. The development of the ORCHID dataset was an effort to address this issue, by pooling data from multiple OPOs. To the best of our knowledge, this is the first study to use ML to analyze organ donor selection factors and multi-transplant in a multi-center OPO setting. The findings highlight the importance of administrative milestones (particularly approaching the donor’s family) and specific organs (heart, lungs) to predicting donor selection and multi-organ transplantation. Overall, given its consistently high recall, precision, and AUC in both research questions, XGBoost (and LR to a smaller extent) has the potential to be adapted into a tool to facilitate equitable and transparent organ allocation in OPOs. However, this study had a few limitations. In its current state, the ORCHID dataset itself was limited; it did not contain comprehensive information about the donors or the administrative processes. In addition, large proportions of MNAR data and multi-collinearity between features prevented the use of the full original ORCHID feature set, which limits the models’ generalizability. This study provides a first step into increasing transparency in OPO procedures, which future studies can expand with more data about the donors and administrative decisions.

## Data Availability

All data produced are available online at https://physionet.org/content/orchid/1.0.0/

## Footnotes

1 https://github.com/chelseatanchip/ORCHID-data-analysis.git

## References

[1] S. D. Shemie et al., “Organ donor management in canada: Recommendations of the forum on medical management to optimize donor organ potential,” CMAJ, vol. 174, no. 6, S13–S30, 2006, ISSN: 0820-3946.

[2] United Network for Organ Sharing. Data and trends., https://unos.org/data/, Accessed Jan 10, 2024.

[3] R. J. Ozminkowski, A. J. White, A. Hassol, and M. Murphy, “Minimizing racial disparity regarding receipt of a cadaver kidney transplant,” American journal of kidney diseases, vol. 30, no. 6, pp. 749–759, 1997.

[4] E. M. Bunnik, “Ethics of allocation of donor organs,” Current Opinion in Organ Transplantation, vol. 28, no. 3, p. 192, 2023.

[5] M. Razdan, H. B. Degenholtz, J. M. Kahn, and J. Driessen, “Breakdown in the organ donation process and its effect on organ availability,” J Transplant, vol. 2015, p. 831 501, 2015.

[6] D. Bertsimas, T. Papalexopoulos, N. Trichakis, Y. Wang, R. Hirose, and P. A. Vagefi, “Balancing efficiency and fairness in liver transplant access: Tradeoff curves for the assessment of organ distribution policies,” Transplantation, vol. 104, no. 5, 2020.

[7] N. Gotlieb et al., “The promise of machine learning applications in solid organ transplantation,” NPJ digital medicine, vol. 5, no. 1, p. 89, 2022.

[8] K. L. Connor, E. D. O’Sullivan, L. P. Marson, S. J. Wigmore, and E. M. Harrison, “The future role of machine learning in clinical transplantation,” Transplantation, vol. 105, no. 4, pp. 723–735, 2021.

[9] H. Adam et al., “Organ retrieval and collection of health information for donation (orchid) (version 1.0.0),” PhysioNet, 2023.

[10] A. Goldberger et al., “Physiobank, physiotoolkit, and physionet: Components of a new research resource for complex physiologic signals,” Circulation, vol. 101, no. 23, e215–e220, 2000.

[11] N. V. Chawla, K. W. Bowyer, L. O. Hall, and W. P. Kegelmeyer, “Smote: Synthetic minority over-sampling technique,” Journal of artificial intelligence research, vol. 16, pp. 321–357, 2002.

[12] E. A. Vail, D. E. Schaubel, P. L. Abt, N. D. Martin, P. P. Reese, and M. D. Neuman, “Organ transplantation outcomes of deceased organ donors in organ procurement Organization-Based recovery facilities versus Acute-Care hospitals,” Prog Transplant, vol. 33, no. 2, pp. 110–120, 2023.

[13] S. M. Lundberg and S.-I. Lee, “A unified approach to interpreting model predictions,” Advances in neural information processing systems, vol. 30, 2017.

[14] C. V. R. Brown et al., “Barriers to obtaining family consent for potential organ donors,” J Trauma, vol. 68, no. 2, pp. 447–451, 2010.

[15] M. Loebe, “Multiple-organ transplantation from a single donor,” Tex Heart Inst J, vol. 38, no. 5, pp. 555–558, 2011.

